# A Streamlined qPCR Method for Characterization of *Enterococcus* spp. Levels in Ambient Surface Water Samples

**DOI:** 10.1101/2025.04.24.25326346

**Authors:** Mano Sivaganesan, Stephanie A. Dean, Jessica R. Willis, Stephanie D. Friedman, Richard Haugland, Orin C. Shanks

## Abstract

Measurement of *Enterococcus* spp. levels with qPCR allows for same-day advisory notification of recreational water quality conditions, representing a major advance over traditional culture-based methods that require 18 or more hours to obtain results. In 2015, the United States Environmental Protection Agency released an *Enterococcus* qPCR protocol for recreational water quality testing. Over the past decade, there have been multiple advances in qPCR-based environmental testing, affording the opportunity to update the current methodology. A streamlined *Enterococcus* qPCR protocol is introduced that simplifies the mathematical model to estimate target sequence concentrations (TSC), reduces sample testing time by 20 min, incorporates a certified control material for standard curve generation, and introduces an inactivated *E. faecalis* whole cell DNA standard (WCDS) control material. A series of experiments were conducted 1) to compare results of the two *Enterococcus* qPCR protocols in analysis of marine, estuarine, and freshwater samples (*n* = 60), 2) to investigate alternative practices to adjust results due to potential water sample matrix interference, control material degradation, and/or analyst inconsistencies, and 3) to evaluate the performance, homogeneity, and stability of an inactivated *E. faecalis* cell preparation as a WCDS control material. Findings indicate a strong correlation between water sample mean log_10_ TSC per reaction results (R^2^ = 0.980) and 100% agreement in amplification and sample processing control tests. A Bayesian approach that accounts for uncertainty in qPCR measurements confirmed statistical equivalence for all water samples yielding paired measurements in the range of quantification, with 72.7% of samples exhibiting reduced error with the new streamlined protocol. Evaluation of three alternative practices to adjust for variation in *Enterococcus* qPCR measurements indicated no significant difference in water sample log_10_ TSC per reaction results with varying concentrations of treated sewage influent. Systematic testing of an inactivated WCDS control material yielded statistically equivalent performance compared to viable *E. faecalis* cell preparations. Homogeneity and stability experiments indicated that *Enterococcus* qPCR measurements of inactivated WCDS are reproducible across multiple preparations and that the material is stable at −20°C for at least 38 weeks. Together, experiments demonstrate that the streamlined protocol and alternative practices should make *Enterococcus* qPCR faster, easier to implement, safer, and more reproducible.

## 1. Introduction

Fecal pollution remains a major challenge for recreational water management worldwide. The most common strategy to identify recreational water conditions is to measure fecal indicator bacteria by cultivation, specifically *Enterococcus* spp. are recommended for the assessment of estuarine and marine waters [1]. Cultivation methods require minimal training and are cost effective; however, they typically require 18 or more hours to yield a result, making same-day beach advisory notification impossible. In addition, studies report that the majority of fecal pollution events occur over 24 h or less time span [2] and delayed water quality information often does not accurately represent current water quality conditions [3, 4]. As a result, rapid fecal indicator bacteria methods are needed that allow for same-day water quality notification. Real-time quantitative PCR methods (qPCR) can yield results in a few hours, representing a potential alternative to more time consuming cultivation approaches [5, 6].

In response, the United States Environmental Protection Agency (USEPA) adapted an *Enterococcus* spp. PCR-based protocol developed for clinical applications [7] to an *Enterococcus* qPCR methodology that can yield quantitative results for recreational water samples in less than 4 hours [8, 9]. Epidemiology studies conducted at fresh and marine coastal beaches predominately impacted by wastewater effluents demonstrated that *Enterococcus* qPCR measurements significantly correlated with gastrointestinal illness occurrence in swimmers [10–12]. This relationship was used to recommend supplemental criteria in the 2012 USEPA Recreational Water Quality Criteria that can be used to issue water quality advisories on a daily basis [13]. Since the release of the *Enterococcus* qPCR method, there is a growing body of evidence demonstrating the benefits of a rapid methodology [3, 4] as well as the public health protection advantages in waters polluted with treated wastewater [14, 15].

Shortly after the conclusion of the epidemiology studies, there were several substantial advances in qPCR-based environmental testing, compelling the modification of the *Enterococcus* qPCR method. For example, the original method was highly prone to amplification inhibition resulting in the need to make dilutions of the DNA extract (1:5 to 1:25) [16], however this practice reduced method sensitivity. The development of a custom DNA polymerase qPCR reagent specifically designed to minimize amplification inhibition from substances commonly found in environmental samples afforded the opportunity to reduce the occurrence of biased measurements and eliminate the need for diluting DNA extracts [17]. In addition, other environmental qPCR-based methodologies started integrating an internal amplification control (IAC) to identify when amplification inhibition is present [18, 19]. While the original *Enterococcus* qPCR methodology included a sample processing control, this practice was unable to differentiate between potential bias in DNA recovery and amplification inhibition, limiting the ability to troubleshoot the cause of biased measurements. Finally, an alternative approach to standardize *Enterococcus* qPCR measurements with epidemiology study results used to derive recreational water quality criteria was identified [20]. These updates to the original *Enterococcus* qPCR protocol were shown to almost eliminate the occurrence of amplification inhibition and improve method reproducibility without significantly influencing mean concentration estimates based on head-to-head testing of ambient freshwater samples [17]. Soon after, the updated *Enterococcus* qPCR methodology was released to the public in 2015 as USEPA Method 1609.1 [21] and has been successfully used in numerous North American coastal beach management scenarios [3, 22, 23].

Since 2015, expanded use and additional scientific advances in qPCR-based environmental testing suggests that a key contributor to the uncertainty in *Enterococcus* qPCR measurements is due to the use of a cell-calibrator [17, 24]. Briefly, the cell-calibrator is used as a positive control where *Enterococcus* and sample processing control qPCR measurements are determined prior to water sample testing (“initial”) and with each batch of water samples, allowing for the adjustment of results based on variability from factors such as cell lysis effectiveness, DNA recovery, qPCR amplification efficiency, and consistency of analyst technique. Modification of the *Enterococcus* qPCR protocol and mathematical model used to estimate *Enterococcus* levels in a water sample could further reduce uncertainty in measurements. In addition, there is greater confidence in the use of shorter qPCR thermal cycling extension times. The *Enterococcus* qPCR method uses a 60 s extension time for the 94 bp 23S rRNA target sequence. Due to the length of the target sequence, it may be feasible to decrease the extension time substantially without compromising method performance. A shorter extension time would reduce the amount of time to generate results, providing water safety information to recreators more quickly.

Another potential area for improvement is the use of a certified standard control material. *Enterococcus* qPCR relies on a standard curve generated from a control material to translate qPCR fluorescence-based measurements into a target sequence concentration [for a review see [25]]. Until recently, there was no certified control material commercially available for the *Enterococcus* qPCR method, so standards had to be prepared by each laboratory, resulting in a considerable amount of measurement variability within and between laboratories [26, 27]. The recent development of a custom certified DNA control material [28] that functions with *Enterococcus* qPCR and 12 other water quality protocols has been shown to generate high-quality standard curves in single [29] and multiple [30] laboratory studies. Finally, the current *Enterococcus* qPCR protocol requires the use of a cell-calibrator preparation consisting of viable *E. faecalis* cells. *E. faecalis* is a gram-positive, commensal bacterium that inhabits the gastrointestinal tract of humans and other animals. It is found in healthy individuals but can be an opportunistic pathogen if it spreads to other parts of the body. As a result, many authorities consider *E. faecalis* to pose a moderate hazard when handling in a laboratory and require specific biosafety precautions (i.e., personal protective equipment, laboratory design, and disposal practices) to minimize exposure [31]. Pivoting from a viable to inactivated *E. faecalis* whole cell DNA standard (WCDS) control material could make implementing the *Enterococcus* qPCR protocol less expensive and safer.

The USEPA recently announced the intention to explore the development of children’s-based recreational water quality risk-based thresholds using *Enterococcus* qPCR [32], providing motivation to revisit the current protocol and update to accommodate for recent advancements. Here, a streamlined *Enterococcus* qPCR protocol is introduced that eliminates a measurement adjustment based on the cell-calibrator, reduces the PCR thermal cycle extension step by 50%, incorporates the National Institute of Standards and Technology Standard Reference Material 2917 (SRM 2917) for standard curve generation, and introduces an inactivated *E. faecalis* WCDS control material. A series of experiments were conducted 1) to determine whether *Enterococcus* qPCR measurements obtained using current [21] and streamlined (this study) methods yield comparable results in ambient surface waters from marine, estuarine, and freshwater sample types exhibiting a wide range of enterococci levels, 2) to investigate alternative sources for sample processing control baseline measurements to identify and adjust results due to potential water sample matrix interference (i.e., biased DNA recovery and/or partial amplification inhibition), and 3) to evaluate the performance, homogeneity, and stability of a commercially available inactivated *E. faecalis* preparation as a WCDS reference material.

## 2. Materials and Methods

### 2.1. Water sample collection and filtration

A total of 124 composite water samples were collected from three sites including a marine beach, an estuary beach, and a freshwater creek situated in Pensacola, Florida (U.S.A.). Sampling occurred from September 2021 to October 2022. Samples were collected in strict accordance with the standard methods for the examination of water and wastewater [33] and transported on ice for culture-based enterococci enumeration and qPCR filtration (< 4 h). Each composite sample consisted of six grab samples (1 L each) arranged along two transects parallel to the respective shoreline corresponding to distances of 0.3 m (3 grab samples) and 1 m (3 grab samples). Transects were arranged in a parallel fashion with each grab sample location situated approximately 50 m apart. For each composite sample, equal sample grab volumes were mixed (total = 6 L) in a sterilized container prior to testing. A field blank consisting of molecular grade water substituted for surface water was included each sampling day (*n* = 42). Water filtration for qPCR testing was conducted on the same composite water samples used for enterococci culture measurements. Briefly, 100 mL of each composite sample was filtered (polycarbonate 47 mm diameter, 0.40 µm pore size; Millipore, Burlington MA, USA), placed in a 2 mL screw cap tube containing silica beads (GeneRite, North Brunswick, NJ USA), and stored at −80 °C (< 18 months).

### 2.2. Enterococcus cultivation and sample selection for head-to-head qPCR testing

*Enterococcus* enumeration was performed for each composite sample within 6 h of sample collection on mEI agar according to Method 1600 [34]. Colonies with blue halos, indicative of enterococci growth, were recorded and expressed as colony forming unit (CFU) per 100 mL. During each sampling event, media sterility checks and negative control using sterile 1X phosphate buffered saline were performed, yielding expected results. *Enterococcus* CFU/100 mL results were used to select 20 samples per site, representing a broad concentration range (**Figure 1**). Approximately 41% of the selected samples (25 of 60) exceed the USEPA Beach Action Value of 1.78 log_10_ CFU/100 mL (60 CFU/100 mL) [13].

**Figure 1.**
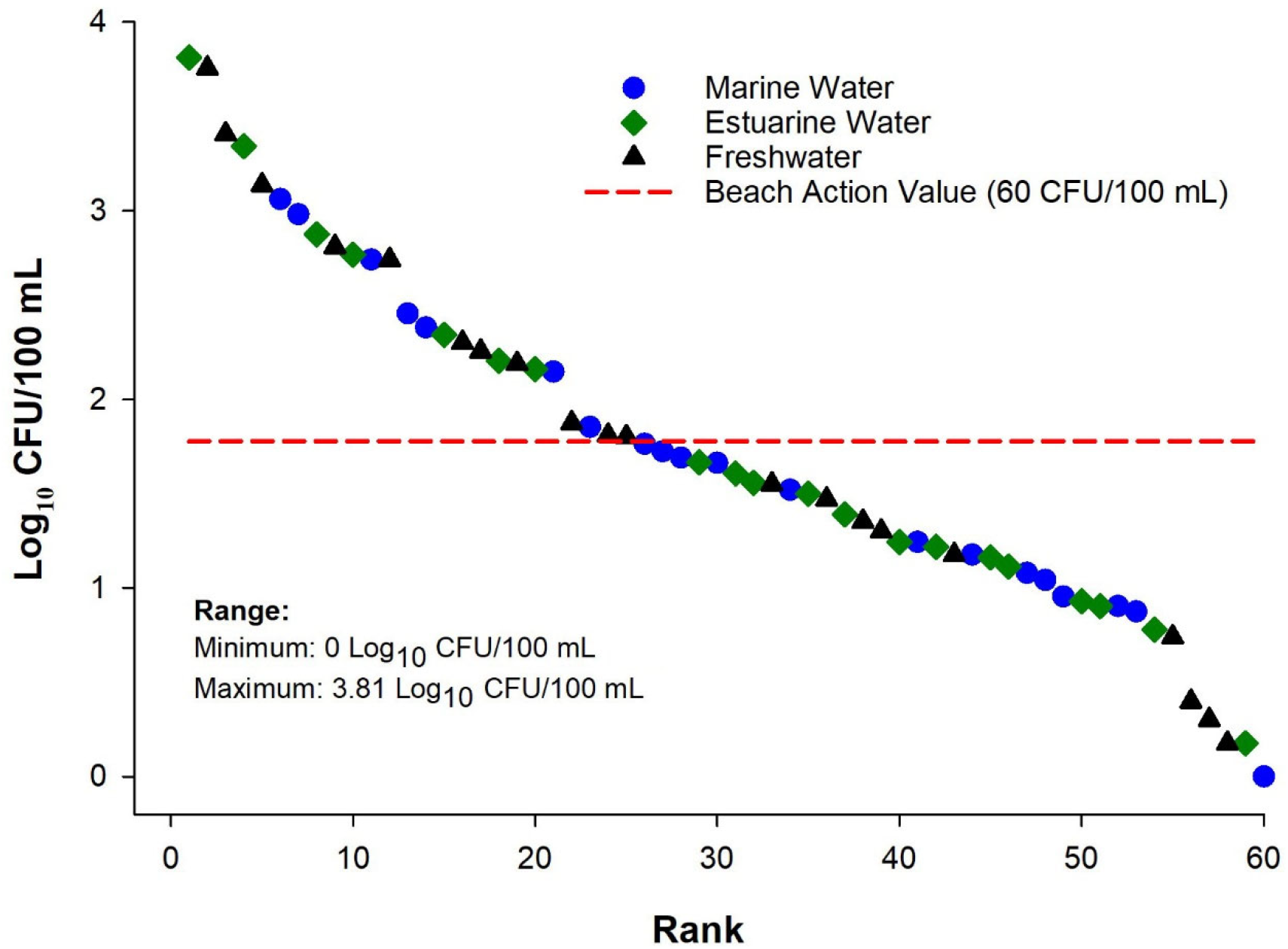
*Enterococcus* log_10_ colony forming unit (CFU) per 100 mL measurements ranked from highest to lowest for marine (circle), estuarine (diamond), and freshwater (triangle) samples selected for head-to-head qPCR testing.

### 2.4. Whole cell DNA standard (WCDS) materials

Whole cell DNA standard (WCDS) materials consisted of *E. faecalis* Multishot 550 BioBall™ (bioMerieux, Lombard, IL; Lot #: 7668, 543.0 ± 43.5 CFU per unit) and inactivated *E. faecalis* cells (Sigma-Aldrich, St. Louis, MO). Cell-calibrator/positive control samples were prepared for extraction with each batch of water samples and for performance assessment experiments. For BioBall™ preparations, each unit was suspended in 1 mL 1X phosphate buffered saline (PBS). For inactivated *E. faecalis* cell preparations, 1 mL of inactivated cells (> 10^8^ bacteria per mL) was diluted with 9 mL of TE buffer (10 mM Tris 0.5 mM EDTA; pH 8), aliquoted into single use 100 µL aliquots, and stored at −20°C. To prepare for each cell-calibrator/positive control extraction, 10 µL of inactivated cell aliquot was mixed with 1 mL of TE buffer. For each filter cell-calibrator/positive control, the entire volume of each WCDS material was filtered (polycarbonate 47 mm diameter, 0.40 µm pore size; Millipore) as described above for water sample filtration.

### 2.5. Enterococcus qPCR reference DNA materials

Reference DNA sources consisted of salmon sperm DNA and two plasmid constructs. A salmon sperm DNA solution containing 10 µg/ml was prepared by diluting a commercially available 10 mg/ml preparation (Sigma-Aldrich, St. Louis, MO). Plasmid constructs included the National Institute of Standards and Technology Standard Reference Material 2917 (SRM 2917; Rockville, MD) for qPCR standard curve generation [28–30] and an internal amplification control (IAC; Integrated DNA Technologies, Coralville, IA). SRM 2917 is a ready to use, commercially available, linearized plasmid consisting of five dilutions ranging from dilution Level 1 (10.3 copies/2 µl) to Level 5 (1.04 · 10^5^ copies/2 µl). The IAC construct was linearized by ScaI-HF restriction digest (New England BioLabs, Beverly, MA), purified via QIAquick PCR Purification Kit (Qiagen, Valencia, CA), quantified with a Qubit dsDNA HS assay kit on a Qubit 3 Fluorometer (Thermo Fisher Scientific, Grand Island, NY), and diluted in 10 mM Tris 0.5 mM EDTA (pH 9.0) to generate 10^2^ copies/2 µL.

### 2.6. DNA extraction

DNA extractions were performed as described in Method 1609.1 [21]. Briefly, AE buffer (Qiagen, Germantown, MD USA) containing 0.2 µg/mL salmon testes DNA, was added to each sample microtube, and subjected to homogenization with a MP FastPrep-24 at 6.0 m/s for 30 s (MP Biomedical, LLC Solon, OH USA). DNA was recovered in the clarified supernatant by centrifugation and directly used as qPCR template (< 30 min holding time at 4 °C). To monitor for extraneous DNA, method blanks consisting of molecular grade water substituted for surface water were prepared in triplicate with each batch of water samples.

### 2.7. qPCR amplification. Enterococcus

qPCR was performed as described in Method 1609.1 [21] using the Entero1a assay [9]. The Sketa22 assay [35] was used as a sample processing control as previously described [21]. All reactions contained 1X TaqMan Environmental Master Mix (version 2.0; Thermo Fisher Scientific), 0.2 mg/ml bovine serum albumin (Sigma Aldrich), 1 µM each primer, 80 nM 6-carboxyfluoroscein (FAM)-labeled probe with TAMRA quencher, and 80 nM VIC/TAMRA-labeled probe. Reactions contained either 5 µL of water sample DNA extract or 2 µL of SRM 2917 in a total reaction volume of 25 µL. Triplicate reactions were performed for all experiments and amplifications were performed on a QuantStudio™ 3 Real-Time PCR System (Applied Biosystems, Foster City, CA USA) with the following thermal cycling profile: 10 min 95°C followed by 40 cycles of 15 s at 95°C and 1 min (current *Enterococcus* qPCR) or 30 s (new streamlined method) at 60°C. Fluorescence thresholds were manually set to 0.03 ΔRN and quantification cycle (Cq) values were exported for further analyses.

### 2.8. qPCR quality controls

A rigorous data acceptance procedure was used to ensure high quality qPCR data. To monitor for potential extraneous DNA contamination during amplification, six no-template controls (NTC) with molecular grade water substituted for template DNA were performed with each instrument run. All individual standard curves were subject to acceptance criteria for linearity (*R*^2^ ≥ 0.980) and amplification efficiency (0.90 to 1.10; *E* = 10^(−1/slope)^ – 1) [36, 37]. The *Enterococcus* duplex IAC protocol was used to assess for potential amplification inhibition in each water sample. Any DNA extract with evidence of amplification inhibition (inhibition threshold = instrument run-specific mean VIC NTC Cq + 1.5 Cq) was discarded from the study [21]. A sample processing control protocol was implemented to monitor for suitable DNA recovery for each water sample as described in Method 1609.1 [21] with the exception that the proposed streamlined *Enterococcus* qPCR protocol uses batch specific Sketa22 qPCR assay measurements only.

### 2.9. Models for Enterococcus

*qPCR target sequence copy (TSC) estimation in an unknown sample*. Method 1609.1 utilizes a ΔΔCq model that incorporates measurements of a cell-calibrator preparation prior to (“initial”) and for each batch of water samples tested. In contrast, the streamlined method relies on a ΔCq model that eliminates the use of “initial” cell-calibrator measurements. Detailed descriptions of each model are provided below.

#### 2.9.1. ΔΔCq Model

The ΔΔCq model used to estimate the number of *Enterococcus* TSC (*X*_*0*_) per reaction volume of a DNA extract from an unknown sample is given by:

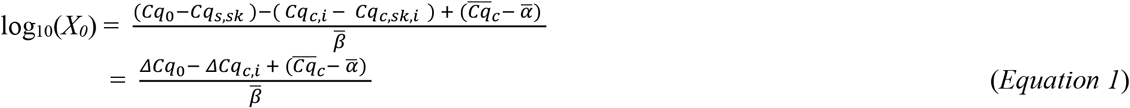

where *ΔCq*_*0*_ (= *Cq*_*O*_ *- Cq*_*s,sk*_) is the difference between the mean Entero1a and mean Sketa22 qPCR assay Cq measurements of each unknown sample, 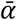 and 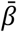 are the “master” standard curve intercept and slope, respectively [26, 38]. *Cq*_*c,i*_ and *Cq*_*c,sk,i*_ are respectively the mean Entero1a and mean Sketa22 qPCR assay Cq values for all cell-calibrator control sample analyses passing the data quality acceptance criteria from the corresponding i^th^ instrument run and their difference is denoted by *ΔCq*_*c,i*_. 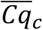 is the mean of all “initial” cell-calibrator sample Entero1a qPCR assay Cq values. *ΔCq*_*0*_, *ΔCq*_*c,i*_, and 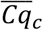 are assumed to have the following known normal distributions:

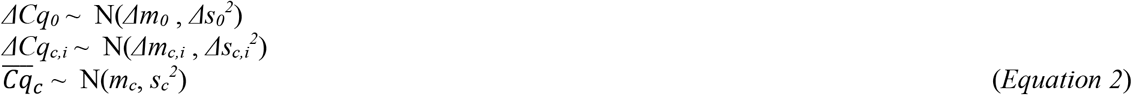

where, *Δm*_*0*_ and *Δs*_*0*_ are the estimated mean and standard deviation of the *ΔCq*_*0*_ values for the three replicate sample filters of each unknown water sample and *Δm*_*c,i*_ and *Δs*_*c,i*_ are the estimated mean and standard deviation of the corresponding cell-calibrator control samples *ΔCq*_*c,i*_ values from the i^th^ instrument run. Within and between filter variabilities were accounted for via a nested analysis of variance (ANOVA) model in estimating *Δs*_*0*_ and *Δs*_*c,i*_. Moreover, the overall mean and standard deviation estimates of the initial cell-calibrators Cq values were used to estimate mean *(m*_*c*_) and standard deviation *(s*_*c*_) of 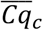. Run-to-run variability, as well as within and between filter variabilities were incorporated in estimating *s*_*c*_ via a nested ANOVA model.

#### 2.9.2 ΔCq Model

When using the ΔCq method to estimate the Entero1a TSC (*X*_*0*_) per reaction in an unknown sample DNA extract, *Equation* (1) is modified to:

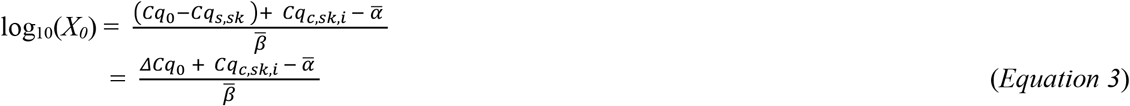

which does not include the ‘initial” or instrument run specific cell-calibrator Cq measurements for the Entero1a assay but does include the mean of the Sketa22 assay Cq measurements (*Cq*_*c,sk,i*_) from the corresponding cell-calibrator/positive control samples in the i^th^ instrument run. *Cq*_*c,sk,i*_ is assumed to have a known normal distribution with mean *m*_*c,sk,i*_ and standard deviation *s*_*c,sk,i*_. As in the case of ΔΔCq model, filter variability was incorporated in *s*_*c,sk,i*_ :

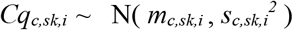

Thus, the ΔΔCq and ΔCq models agree, in theory, if the mean Entero1a assay Cq values of the run specific cell-calibrator/positive control samples and the “initial” cell-calibrator samples are the same, i.e.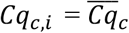.

### 2.10. Additional data analyses

Standard curves were generated from six independent runs and combined to generate a ‘master’ curve using a Bayesian Markov Chain Monte Carlo approach[26, 38]. Outliers were defined as the absolute value of a studentized residual > 3. The lower limit of quantification (LLOQ) was defined as the upper bound of the 95% Bayesian confidence interval of the estimated Cq at the lowest DNA standard concentration (SRM 2917, 10.8 copies per reaction) used to generate individual standard curves. Repeated measures from a water sample (3 filters . 3 replicates per filter = 9 total reactions) were eligible for TSC concentration determination if six or more individual reactions exceeded the respective LLOQ (66.7% of total reactions). Water sample log_10_ TSC per reaction with 95% Bayesian credible intervals (95% BCI) generated from ΔΔCq and ΔCq model protocols were compared using a Bayesian approach that accounts for uncertainty in the respective “master” curve, sample qPCR measurements, and filter-to-filter variability. To compare mean log_10_ TSC per reaction paired measurements, a Pearson product momentum correlation analysis (α = 0.05) was used.

To evaluate the homogeneity of vendor inactivated *E. faecalis* whole cell DNA standard (WCDS) materials, six commercial stock preparations were obtained and systematically tested with the Entero1a assay as follows: the first tube was subject to six instrument runs with each run consisting of three filters with triplicate measurements (3 filters · 3 replicates/filter x 6 instrument runs = 54 measurements), then tubes two through six were tested with a single instrument run (3 filters · 3 replicates/filter = 9 measurements). Run-to-run variability, as well as within and between filter variabilities were incorporated in estimating the standard deviation for the first tube via a nested ANOVA model. An Entero1a mean Cq with 95% confidence interval (CI) was then used to establish acceptance metrics to evaluate homogeneity of tubes two through six. A similar experimental design was used to evaluate stability, where time zero (T_0_ = stored at −20°C for 24 h) aliquots were tested across six instrument runs to calculate an Entero1a mean Cq with 95% CI and standard deviation to elevate additional time points (T_1_ = 1 week, T_2_ = 6 weeks, T_3_ = 12 weeks, and T_4_ = 38 weeks) based on a single instrument run. All statistics were calculated with SAS software (Cary, NC) or WinBugs (https://www.mrc-bsu.cam.ac.uk/software/bugs/thebugs-project-winbugs).

## 3. Results

### 3.1. qPCR quality controls

High-quality data for qPCR experiments were identified using a series of data acceptance metrics and controls. Across all standard curves, a total of five outliers (2.8%) were observed (180 total measurements) with all instances corresponding to the lowest concentration used as template (SRM 2917 Level 1 = 10.3 copies/2 µl). Individual standard curves generated from the SRM 2917 control material exhibited amplification efficiencies (*E*) ranging between 0.94 to 1.02 and R^2^ values ≥ 0.995, regardless of model protocol. Extraneous DNA control reactions (NTC and method blanks) indicated the absence of contamination in 95.8% reactions (1,059 total reactions). All false positives yielded Cq values greater than LLOQ (33.1 Cq for ΔΔCq and 32.9 Cq for ΔCq protocols). Evidence of amplification inhibition was absent in all water sample experiments. Instrument run specific amplification inhibition Cq thresholds ranged from 32.9 to 34.7 for ΔΔCq and 33.0 to 34.5 for ΔCq protocols. A single DNA extract failed SPC testing (1.7%; 1 of 60 DNA extracts) with both protocols identifying the same sample. Sketa22 qPCR cell-calibrator/positive control Cq standard deviations were ≤ 0.59 (both protocols). “Initial” cell-calibrator (*E. faecalis* Multishot 550 BioBall™) mean Cq and standard deviations were generated from repeated Entero1a (31.00 ± 0.38) and Sketa22 (18.52 ± 0.30) qPCR measurements across six instrument runs for the ΔΔCq protocol.

### 3.2. ‘Master’ standard curve comparison

Both protocols utilize a ‘master’ standard curve generated from six independent instrument runs where the qPCR extension step is either 60 s (ΔΔCq) or 30 s (ΔCq). ‘Master’ intercepts were 36.5 ± 0.08 (ΔΔCq) and 36.2 ± 0.08 (ΔCq). ‘Master’ slopes included −3.36 ± 0.02 (ΔΔCq) and −3.35 ± 0.02 (ΔCq) corresponding to *E* values of 0.99 for both protocols. A Bayesian comparison of ‘master’ slope values (95% BCI lower bound = −0.05; upper bound = 0.05) indicated no significant difference (ratio 95% Bayesian credible interval intersects 0), while ‘master’ intercept values (95% BCI lower bound = 0.06; upper bound = 0.49) were significantly different (ratio 95% Bayesian credible interval did not intersect 0).

### 3.3. Comparison of water sample Enterococcus qPCR concentration estimates

To evaluate the performance of the new streamlined ΔCq protocol, 60 water samples collected from multiple water types, exhibiting a wide range of culture-based enterococci levels (**Figure 1**), were compared in head-to-head experiments. Paired measurements indicated 93.3% (56 of 60 samples) agreement in eligibility assignment for concentration determination (criteria: six or more sample Cq measurements in the range of quantification). For all instances of disagreement, individual Cq measurements were within 0.5 Cq (on average) of the respective LLOQ, indicating that the magnitude of disagreement was minor. A total of 33 samples yielded paired mean log_10_ TSC per reaction with 95% BCI concentration estimates (**Figure 2, Panel A**). To determine if protocol concentration estimates were significantly different for a given sample, a mean log_10_ ratio with 95% BCI was determined for each sample (**Figure 2, Panel B**). No significant difference was observed between sample paired measurements (log_10_ ratio 95% BCI intersects 0). In addition, Pearson’s correlation analysis of mean log_10_ TSC per reaction paired estimates yielded an R^2^ = 0.980 (p < 0.0001). While concentration estimates were highly correlated and not significantly different, 72.7% (24 of 33) of samples exhibited smaller standard deviations with the new streamlined ΔCq protocol. Percent reduction in error [(ΔΔCq standard deviation – ΔCq standard deviation) / ΔCq standard deviation] for these samples ranged from 2.2% to 73.5% with an average of 31.6%.

**Figure 2.**
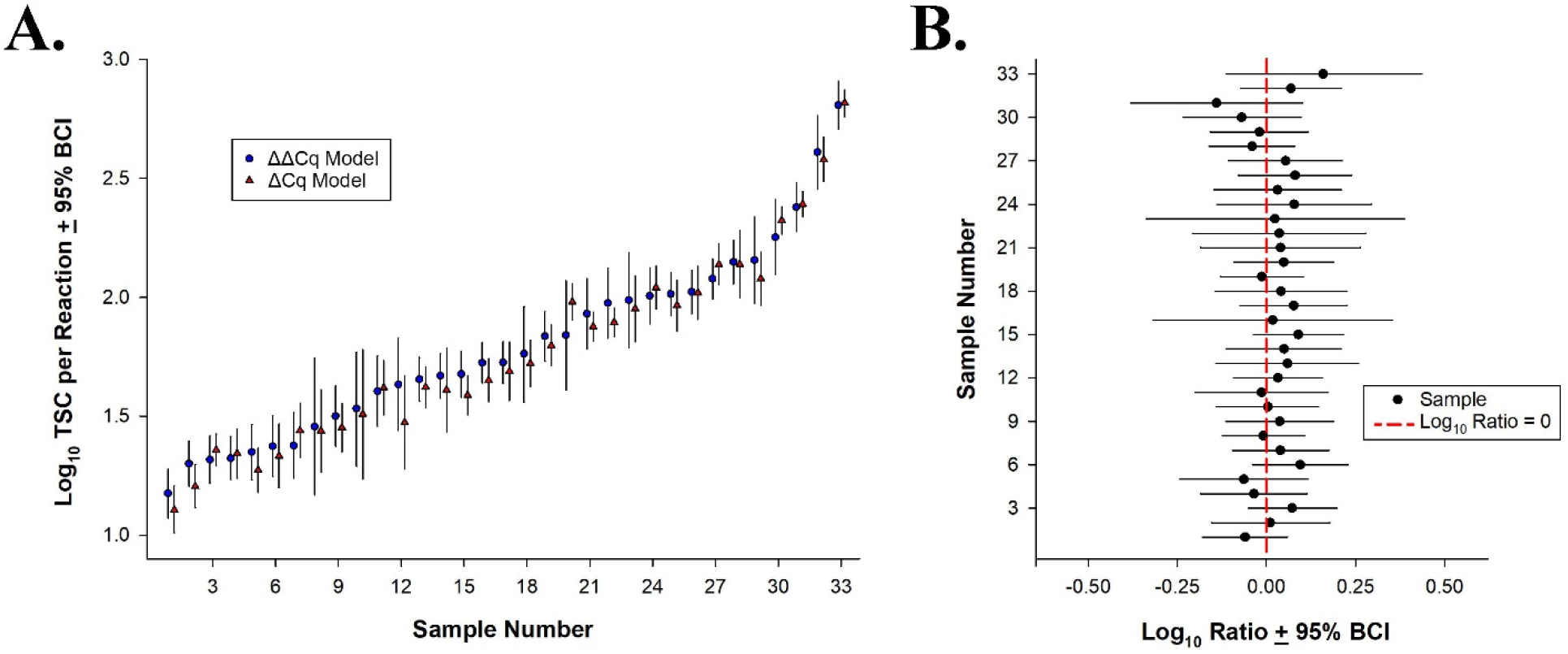
Comparison of mean log_10_ target sequence copies (TSC) per reaction with 95% Bayesian Credible Interval (95% BCI) estimates by sample using ΔΔCq (circle) and new streamlined ΔCq (triangle) protocols (**Panel A**) and sample corresponding mean log_10_ ratios with 95% BCI (**Panel B**). Error bars depict respective 95% BCI ranges. The vertical red line in **Panel B** depicts a log_10_ ratio of zero. Mean log_10_ ratio values with a 95% BCI that intersect the vertical red line indicate no significant difference between ΔΔCq and ΔCq protocol estimates.

### 3.4. Alternative sources of Sketa22 measurements for the streamlined ΔCq protocol

The streamlined ΔCq protocol includes a mandatory Entero1a Cq adjustment based on the difference in Sketa22 Cq measurements from an instrument run specific positive control and the unknown water sample. Using two water samples containing different concentrations of treated sewage effluent, *Enterococcus* qPCR log_10_ TSC per reation with 95% BCI were estimated using WCDS materials (BioBall^®^ or inactivated *E. faecalis* cells) and method blanks (1X PBS or laboratory grade water) as the source of instrument run-specific Sketa22 qPCR measurements (**Figure 3, Panel A**). Mean log_10_ ratios with 95% BCI (**Figure 3, Panel B**) indicate no significant difference (95% BCI does not intersect 0) for all Sketa22 qPCR source combinations (six possible combinations).

**Figure 3.**
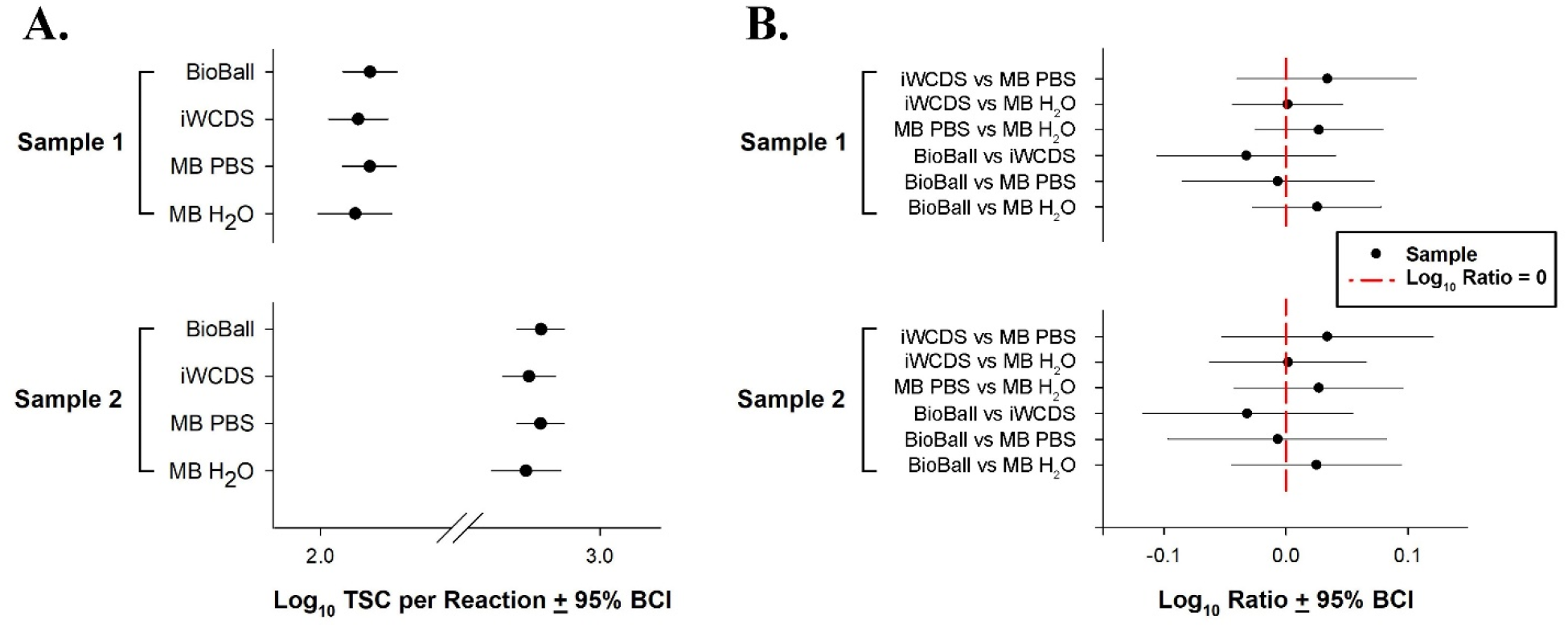
Comparison of mean log_10_ target sequence copies (TSC) per reaction with 95% Bayesian Credible Interval (95% BCI) estimates for two water samples polluted with different concentrations of treated sewage effluent using four Sketa22 qPCR measurement sources (**Panel A**) and sample corresponding mean log_10_ ratios with 95% BCI for each possible Sketa22 qPCR measurement source combination (**Panel B**). Sketa22 qPCR measurement sources include BioBall (BioBall^®^ whole cell DNA standard), iWCDS (inactivated Whole Cell DNA Standard), and MB PBS (method blank with 1X PBS), and MB H_2_O (method blank with laboratory grade water). The vertical red line in **Panel B** depicts a log_10_ ratio of zero. Mean log_10_ ratio values with a 95% BCI that intersect the vertical red line indicate no significant difference between water sample concentration estimates.

### 3.5. Performance, homogeneity, and stability of inactivated whole cell DNA standard (WCDS) material

An ideal WCDS reference material should yield consistent results across a wide range of concentrations, exhibit homogeneity from one vendor stock preparation to another, and be stable during storage for a known time. To evaluate *Enterococcus* qPCR measurement precision of the inactivated WCDS, repeated measures (three filters with triplicate measurements per filter) at four 10-fold dilutions were treated as an unknown sample, allowing for mean log_10_ TSC per reaction with 95% BCI and standard deviation determination. Mean log_10_ TSC with 95% (lower bound, upper bound) concentration estimates ranged from 4.81 (4.71, 4.90) to 1.74 (1.69, 1.79) with standard deviations ≤ 0.05. To assess vendor stock homogeneity, six commercially obtained stocks were systematically tested, indicating that all individual Entero1a Cq measurements from tubes two to six fall within the mean Cq 95% CI acceptance range determined from repeated measures of tube one (**Figure 4, Panel A**). To establish a shelf-life under −20°C storage conditions, aliquots of an inactivated WCDS preparation were systematically tested at six time points (T_0_ to T_5_) to establish metrics that account for Entero1a Cq variability within and between instrument runs (**Figure 4, Panel B**). Stability experiments indicate that the inactivated WCDS material yields similar Cq measurements (all individual Cq measurements within 95% CI) for at least 38 weeks (all time points tested).

**Figure 4.**
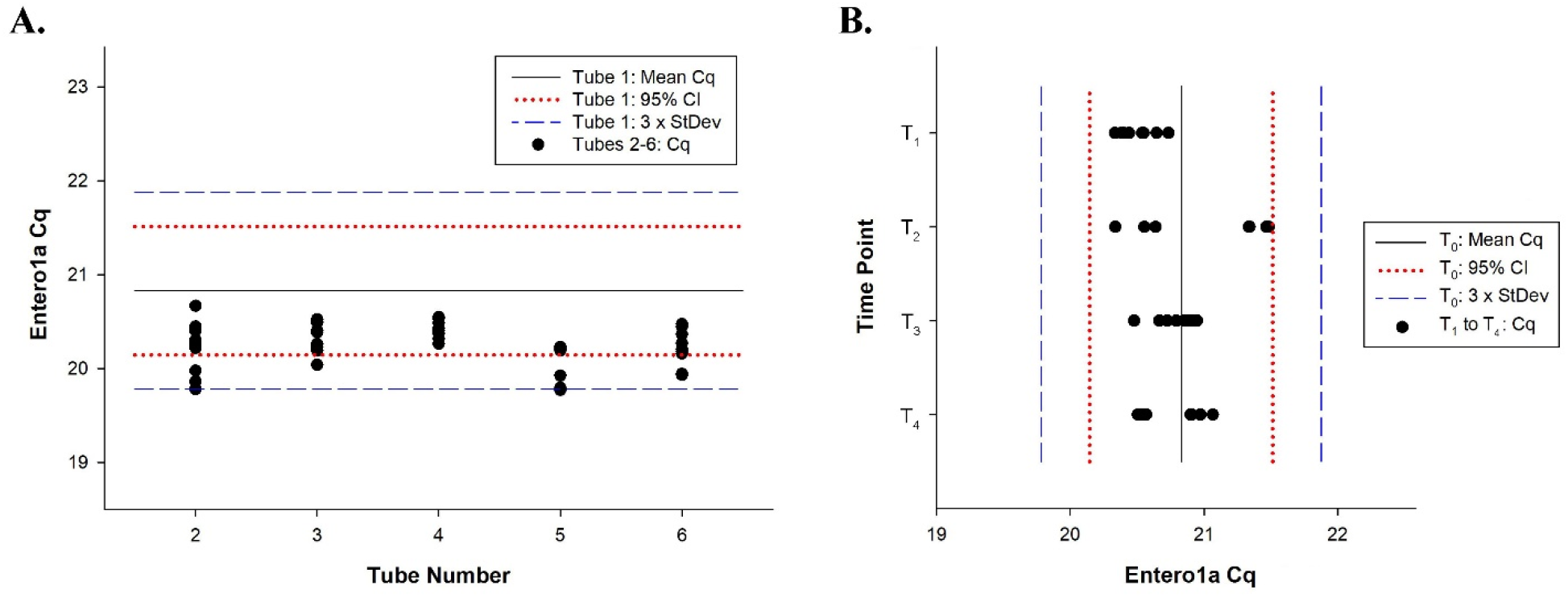
Evaluation of inactivated *E. faecalis* whole cell DNA standard (WCDS) material homogeneity (**Panel A**) and stability at −20°C (**Panel B**). Black circles denote individual Entero1a Cq measurements. The widest interval, indicated by the dashed lines represent Entero1a Cq standard deviation intervals (3 · standard deviation) based on repeated measures across six instrument runs. The dotted lines corresponds to respective 95% Confidence Interval (95% CI) ranges. Time points include T_0_ = stored at −20°C for 24 h, T_1_ = 1 week, T_2_ = 6 weeks, T_3_ = 12 weeks, and T_4_ = 38 weeks.

## 4. Discussion

### 4.1. Streamlined ΔCq protocol yields similar concentration estimates

Head-to-head testing of surface waters exhibiting a wide range of cultivated *Enterococcus* concentrations (CFU/100 mL) and water compositions was conducted to compare ΔΔCq and ΔCq protocols. A total of 33 samples yielded paired measurements in the range of quantification exhibiting a remarkable level of agreement (R^2^ = 0.980). Furthermore, a Bayesian-based approach accounting for uncertainty in ‘master’ standard curves, sample qPCR measurements, and filter-to-filter variability further confirmed statistical similarity (**Figure 2**). Both protocols showed 100% agreement for amplification inhibition and sample processing control tests, demonstrating that each procedure identified matrix interference in an equivalent fashion and that measurement bias from environmental factors was rare in subtropical fresh, estuarine, and marine water types tested in this study. Strong agreement was expected because both protocols utilize identical primer and probe sequences, qPCR reagents, qPCR thermal cycling profile (except extension step), and reference DNA materials. In addition, both methods use the same filtration and DNA recovery protocols. Together findings provide robust evidence that the new streamlined ΔCq protocol that reduces the qPCR extension step by 50% and eliminates a measurement adjustment based on a cell-calibrator should yield highly equivalent TSC concentrations to the established, nationally validated *Enterococcus* qPCR method (ΔΔCq protocol).

### 4.2. Benefits and challenges of eliminating the Enterococcus qPCR cell-calibrator measurement adjustment

In the ΔΔCq protocol, “initial” and instrument run specific cell-calibrator Entero1a qPCR measurements are used to adjust corresponding Entero1a water sample Cq measurements (*Equations 1 and 2*). However, studies have demonstrated that this adjustment can be a source of error [17, 24]. The elimination of this adjustment practice in the streamlined ΔCq protocol has multiple benefits. First, the new protocol resulted in lower measurement error in most water samples with standard deviation percent reductions as high as 73.5%, suggesting that the streamlined protocol should allow for more reproducible *Enterococcus* qPCR measurements within and between laboratories. A less obvious, but important benefit is the elimination of variability due to analyst method proficiency and/or potential cell-calibrator preparation degradation between “initial” and sample batch testing. In the ΔΔCq protocol, repeated measures of a cell-calibrator preparation are conducted prior to water sample testing (“initial”) and then used to establish acceptance metrics (acceptance threshold = “initial” mean Cq ± 3 . standard deviation). Cell-calibrator measurements from the same preparation are then included in subsequent water sample test batches, often over an entire beach season (≥ 15 weeks) and must fall within the established “initial” acceptance range to be eligible for concentration estimation. While a defined cell-calibrator acceptance range limits the amount of variability between “initial” and batch specific measurements, the maximum allowable standard deviation to establish these metrics is 1 Cq [21], allowing for Entero1a and Sketa22 Cq adjustments as large as 3 Cq (acceptance range = 3 . 1 Cq standard deviation). As a result, a comparison of the same water sample from one batch to another could result in substantially different concentration estimates due to inconsistent method proficiency and/or partial degradation of the cell-calibrator preparation (~1 order of magnitude range; 3.3 Cq shift represents a theoretical 10-fold concentration shift).

While there are advantages for the ΔCq protocol, it does create some challenges. Namely, there is a need to establish a sample processing method proficiency metric to ensure a practitioner is wielding the method with acceptable reproducibility. In this single laboratory study, the ΔCq protocol Sketa22 qPCR standard deviation averaged 0.29 ± 0.11 Cq across 11 sample batches using the BioBall^®^ WCDS as a positive control (range: 0.12 Cq to 0.58 Cq). While this value could serve as a method proficiency metric for future practitioners, an interlaboratory study would be ideal to identify a standard deviation that incorporates lab-to-lab variability like ones reported for other qPCR recreational water quality qPCR standardized methods [39].

### 4.3. Inactivated E. faecalis cells as an alternative positive control material

An *E. faecalis* WCDS material can be a reliable source of instrument run specific Sketa22 measurements under ideal conditions as well as serve as a positive control to ensure adequate DNA recovery from filtered cells. Historically, WCDS materials consist of viable cells either prepared by a laboratory prior to water sample testing or commercially obtained (e.g., BioMerieux *E. faecalis* BioBall^®^). However, lab prepared materials require a laboratory equipped to cultivate, enumerate, and store viable cell stocks. Commercially available BioBall^®^ preparations are certified by the International Organization for Standardization for CFU ± 10% based on flow cytometry measurements and do not require lab cultivation or enumeration. However, both options require handling of viable E. *faecalis* cells which are considered by some authorities to be a moderate safety risk, requiring specialized laboratory facilities, equipment, and personal protective equipment [31]. An alternative to viable cell materials is a commercially available inactivated *E. faecalis* cell preparation which would eliminate the potential need for additional safety practices. Findings demonstrate that inactivated WCDS dilutions representing four orders of magnitude yield extremely consistent *Enterococcus* qPCR measurements (mean log_10_ TSC standard deviations ≤ 0.05), demonstrating that this material can be measured with a high level of confidence.

To further explore the suitability of inactivated WCDS preparations for an *Enterococcus* qPCR application, a head-to-head experiment was conducted where Sketa22 qPCR measurements from viable (BioBall^®^) and inactivated WCDS preparations were used to estimate *Enterococcus* levels in waters polluted with treated sewage at two concentrations (**Figure 3**). Findings indicate statistically similar results (ratio 95% BCI intersects 0), demonstrating that the inactivated WCDS material can be substituted for viable cell preparations without compromising method performance. To further validate the inactivated WCDS material, systematic testing of multiple vendor stocks was conducted to evaluate homogeneity. Inactivated WCDS aliquots were also tested to establish a shelf-life expectancy when stored at −20°C. Findings indicate similar *Enterococcus* qPCR measurements across vendor stocks and that inactivated cell preparation aliquots are stable at −20°C for at least 38 weeks (**Figure 4**), suggesting that the inactivated *E. faecalis* WCDS material should be a reliable substitute for viable cell materials.

### 4.4. The elimination of whole cell DNA standard (WCDS) from routine Enterococcus qPCR testing

The streamlined ΔCq protocol utilizes instrument run specific repeated Sketa22 qPCR measurements from a WCDS positive control to compare sample processing under ideal conditions to those in a water sample. If Sketa22 qPCR measurements from the positive control and water sample are identical, then no Entero1a qPCR measurement adjustment is necessary. However, if there is a difference, a shift in the corresponding mean Entero1a qPCR measurement from the water sample is made (see *Equation* 3) to account for potential DNA recovery bias, partial amplification inhibition, and/or variability in analyst technique. As a result, the ability to generate consistent Sketa22 measurements with each sample batch under ideal conditions is critical. While using a WCDS positive control (i.e., cell-calibrator) has been traditionally used for *Enterococcus* qPCR applications, using instrument run specific Sketa22 measurements from an alternative source (method blank) and eliminating the use of a WCDS material from routine *Enterococcus* qPCR testing could have several advantages. For example, handling high concentrations of *E. faecalis* cells in the laboratory environment on a regular basis, where trace analysis (≥ 40 qPCR cycles) of a multiple copy per genome target sequence (23S rRNA gene) is conducted, increases the risk of cross-contaminating samples and laboratory workspaces. While extraneous DNA controls demonstrated the absence of contamination in 95.8% of test reactions in this study, it is possible that the exclusion of WCDS materials in experiments may have eliminated the occurrence of any false positives. The omission of WCDS materials would also further reduce the cost of conducting *Enterococcus* qPCR on a routine basis.

Experiments comparing the influence of Sketa22 qPCR measurements orginating from WCDS materials (BioBall^®^ and inactivated *E. faecalis* cells) with those from method blanks (laboratory grade water and 1X PBS) demonstrate concentration estimate statistical equivalence in sewage polluted water samples (**Figure 3**), providing an alternative to the inclusion of a WCDS-based control with each sample test batch. The elimination of WCDS materials on a regular basis is a common practice for other standardized water quality PCR-based protocols [39–41], including an *Enterococcus* digital PCR method reported to generate statistically similar results compared to the ΔΔCq protocol [42]. However, it would still be useful for practitioners to demonstrate method proficiency with a WCDS material on an episodic basis. For example, during method training and when key changes are made to laboratory workflow (i.e., new analyst, updated instrumentation, new reagent lot). This could be accomplished by developing a method proficiency test protocol that incorporates a WCDS positive control, such as inactivated *E. faecalis* preparations.

## 5. Conclusions

A streamlined ΔCq protocol for *Enterococcus* enumeration is introduced that yields statistically equivalent concentration estimates to a nationally validated protocol (ΔΔCq). The new method includes a modified qPCR extension step that reduces recreational water sample processing time by 20 min, allowing for more rapid notification of water quality and improving public health protection. In addition, most water sample concentration estimates exhibited a reduced error compared to the current ΔΔCq protocol, which should lead to more reproducible results within and between laboratories for future practitioners. An alternative WCDS material consisting of inactivated *E. faecalis* cells was also introduced and validated for use in *Enterococcus* qPCR applications either on a routine basis as a positive control or episodically as a total workflow control to evaluate laboratory method proficiency. An inactivated WCDS material eliminates the need for additional safety precautions often required when handling viable *E. faecalis* cell preparations, making method implementation less complex and safer. Finally, an alternative source of sample processing control measurements (Sketa22 qPCR) used to adjust for potential bias in sample measurements due to matrix interferences and/or analyst inconsistencies is proposed that can eliminate the use of WCDS materials from routine *Enterococcus* qPCR testing. However, an interlaboratory assessment of the streamlined *Enterococcus* qPCR protocol (ΔCq) to establish a method proficiency protocol with custom metrics that account for lab-to-lab variability to help ensure suitable method implementation would be beneficial for large-scale implementation.

## Data Availability

All data produced in the present study are available upon reasonable request to the authors

## Acknowledgements

Information has been subjected to U.S. EPA peer and administrative review and has been approved for external publication. Any opinions expressed in this paper are those of the authors and do not necessarily reflect the official positions and policies of the U.S. EPA. Any mention of trade names or commercial products does not constitute endorsement or recommendation for use.

